# Intrinsic and extrinsic connectivity of the seizure onset zone at rest and during stimulation

**DOI:** 10.64898/2026.02.27.26347224

**Authors:** Joshua J. LaRocque, William K.S. Ojemann, Jay Xu, Alfredo Lucas, Nishant Sinha, Eli J. Cornblath, Caren Armstrong, Samuel Tomlinson, Eric Marsh, Saurabh R. Sinha, Brian Litt, Kathryn Davis, Quy Cao, Erin C. Conrad

## Abstract

About half of patients who undergo epilepsy surgery for drug-resistant epilepsy have seizure recurrence, supporting the need for approaches that more accurately identify the *epileptogenic zone*, defined as the brain areas whose removal causes cessation of seizures. Altered network connectivity has emerged as a candidate biomarker of the epileptogenic zone, but how connectivity is altered in the epileptogenic zone remains uncertain, with prior studies reporting inconsistent results. We hypothesized that a difference in intrinsic versus extrinsic connectivity of the epileptogenic zone may explain prior discrepant findings. We studied a multicenter cohort of adult and pediatric patients who underwent intracranial EEG recording and brain stimulation as part of epilepsy surgery planning. We measured spontaneous connectivity using Pearson correlation and perturbational connectivity using stimulation evoked potentials, modeling the connectivity according to the location of contacts in relation to the seizure onset zone (SOZ) while controlling for inter-electrode distance. We analyzed 79 patients (37 adults, 42 children). For both adult and pediatric patients, resting connectivity was higher within compared to outside the SOZ, but resting connectivity between SOZ and non-SOZ contacts was reduced. Stimulation connectivity followed a similar pattern, with elevated within-SOZ connectivity but reduced connectivity between SOZ and non-SOZ. The results support the hypothesis that the epileptogenic zone is disconnected from the rest of the brain but intrinsically hyperconnected. This result helps reconcile prior inconsistencies across studies, aligns with the results of basic science studies, and suggests that future translational work should model this heterogeneous pattern to increase the yield of using connectivity to localize the epileptogenic zone.

## Introduction

Roughly one-third of people with epilepsy continue to have seizures despite antiseizure medications.^1^ For these patients, surgical resection of brain tissue generating seizures – the epileptogenic zone (EZ)^1^ — offers the best chance of cure. Identifying the EZ is difficult; many patients undergo invasive intracranial EEG (iEEG) monitoring for 1–2 weeks to localize the seizure onset zone (SOZ). Despite this lengthy and resource-intensive evaluation, about half of surgical patients experience seizure recurrence after surgery.^2,3^ We need more accurate, less morbid, and less expensive methods to identify the EZ and to characterize the associated networks.

Concurrent with the increasing recognition of epilepsy as a network disorder, interictal (between seizures) network connectivity has emerged as a candidate biomarker that could improve and shorten presurgical evaluation. However, findings are inconsistent: some studies report increased connectivity within the EZ, consistent with rapid recruitment during seizures.^4^ Other studies of diffusion tractography,^5^ resting state fMRI,^6,7^ and resting intracranial EEG^8^ reported reduced connectivity in the EZ. Other work suggests more nuanced patterns—differences within SOZ regions versus between SOZ and non-SOZ regions, and asymmetries in connectivity to versus from the SOZ.^9,10^

Electrical brain stimulation is routinely used during surgical planning to evoke seizures, probe eloquent cortex, and interrogate network architectures. Stimulation provides a complementary, causal probe of network organization, revealing connections in response to a perturbation. However, stimulation studies also diverge, with reports of both heightened^11–14^ and diminished^15^ responses in SOZ tissue.

These inconsistent findings limit our understanding of epileptic pathology and our ability to use network alterations as a biomarker to guide surgical planning. Potential explanations for these discordant results include: 1) most studies include small, heterogeneous cohorts; 2) there is inconsistent application of controls for inter-electrode distance, a known confound in connectivity measures,^8,16^ and 3) it is possible that network alterations in the EZ are too complex to be summarized as merely “increased vs decreased.”

To address these gaps, we examine connectivity in two complementary modalities—baseline interictal iEEG functional connectivity and stimulation-evoked connectivity—in adults and children with drug-resistant epilepsy undergoing iEEG monitoring at two epilepsy centers. We compare undirected and directed connectivity to and from SOZ regions, testing the hypothesis that the EZ demonstrates altered connectivity, and that there are discrepant patterns for intrinsic vs. extrinsic EZ connectivity.

## Methods

### Patient population

37 adult and 42 pediatric patients were included in this study. Subjects were patients with epilepsy undergoing intracranial EEG at the Hospital of the University of Pennsylvania (HUP) and the Children’s Hospital of Philadelphia (CHOP),^17^ respectively, from 2019-2024 who underwent low-frequency electrical stimulation at a sufficient number of sites for analyses to be performed, and who had imaging data available to perform electrode reconstructions. Stimulation was performed under IRB-approved research (N=11 HUP patients; all CHOP patients) or clinical protocols (HUP, N = 26), with IRB-approved retrospective analysis of the latter cohort.

### Clinical data

SOZ sites were determined by the clinical team caring for the patient, as recorded in the EEG report in the electronic medical record. This is generally determined by identifying the earliest changes thought to represent a definite ictal pattern as the SOZ. Epilepsy localization was determined by the clinical team and confirmed in a surgical case conference, and typically based on consistent SOZ, with imaging and other clinical data informing the decision.

### Recording and stimulation procedure

Electrode placement was determined by the clinicians (neurologists and neurosurgeons) who were involved in the patients’ care, informed by an epilepsy surgery conference discussion. For adult recordings, depth electrodes contained 4–12 contacts, each 1.10 mm in diameter and 2.41 mm in length, spaced 5 mm apart (center-center) along the shaft. For pediatric recordings, electrodes contained 5–18 contacts, each 0.8 mm in diameter and 2 mm in length, with 3.5 mm center-center spacing between adjacent contacts. In some longer electrodes used in pediatric recordings, there were more widely spaced contacts (10-12 mm), which were not used for stimulation. In the adult patients, electrical stimulation typically occurred the day after the surgical implantation of electrodes, whereas for the pediatric patients it was typically delayed until the end of the admission. In nearly all occasions, patients were on home or near-home doses of anti-seizure medications. During recording, signals were referenced to an electrode contact chosen by clinicians that was distant from the suspected seizure onset area, typically in bone or in remote white matter. Regarding stimulation strategy, the goal was to stimulate every adjacent pair of electrode contacts definitively located within the brain parenchyma (omitting those contacting dura).

Electrodes suspected to be distant from the SOZ were sometimes not stimulated due to time constraints. Stimulation in an anatomical region was stopped if a seizure or noxious clinical response was provoked. Patients at HUP received bipolar, biphasic stimulation with the following parameters: 1 Hz, 300-500 μs pulse width (300 μs for the first 11 patients and 500 μs thereafter), 3 mA amplitude (charge density 11.1 or 18.5 μC/cm). Patients at CHOP had variable stimulating parameters, with amplitudes ranging from 1-8 mA, frequencies ranging from 1-2 Hz, and pulse widths ranging from 300-500 μs (charge density 17.9-59.7 μC/cm^2^). The cathode was typically designated as the most superficial and the anode as the deepest contact for the initial pulse phase.

### Analysis: Electrode Localization

To map the electrode contacts from the post-implant MRI scan to the pre-implant T1-weighted MRI image, we utilized iEEG-recon^18^ for the adult data and EpiTools^19^ for the pediatric data. The electrode contacts were assigned to anatomical regions from the Desikan-Killaney atlas based on a 5 mm sphere surrounding the electrode, following the methods outlined previously^18^; this information was used to identify each electrode contact as being within gray or white matter. A bipolar pair of electrode contacts was collectively considered to be gray matter if either of the contacts resided within gray matter. Bipolar contact pairs, or channels, were considered to be in white matter if at least one contact of the pair resided within white matter, and neither were in gray matter. The reconstruction also yielded 3-dimensional coordinates for each electrode contact, which were used to calculate pairwise distances between all bipolar channels for each subject. The midpoint of bipolar channels was used for measuring inter-channel distances.

### Analysis: Data processing

The data were analyzed within Matlab using custom scripts (available on https://github.com/erinconrad/CCEPS), as described previously.^15^ All electrographic signals (recorded in microvolts) were converted to a bipolar montage as is commonly done in clinical and research settings. This was done by taking the difference in the signals between each adjacent pair of contacts along the electrode shaft, thereby creating bipolar channels. Contacts designated as EKG, scalp electrodes, or EMG were not analyzed. The annotations generated in the EEG by the Natus stimulator were used to identify time periods of stimulation.

### Analysis: Undirected connectivity from spontaneous activity

We performed parallel analyses in both adult and pediatric data sets to remain sensitive to distinct patterns of results in the two patient groups (Table 1). To understand how connectivity derived from spontaneous neural activity is related to the SOZ, we computed pairwise correlations between the spontaneous activity recorded from all bipolar channels located within gray matter. To do this, we identified a 60-second time period immediately prior to beginning stimulation, filtered the signals between 0.5 and 55 Hz, and computed time series correlations for each one-second time window. We then calculated a mean correlation across these time windows for each pair of bipolar derivations to obtain a single correlation value per pair of bipolar derivations. Finally, we took the absolute value of these correlations, because the significance of positive vs negative correlations in a bipolar montage depends on the arbitrary order of subtraction. We restricted our primary baseline connectivity analyses to gray matter (GM) sites only. This was because we anticipated that most SOZ sites would be located within GM, and we wished to avoid any potential for this to bias our results comparing SOZ to non-SOZ responses.

**Table 1:**
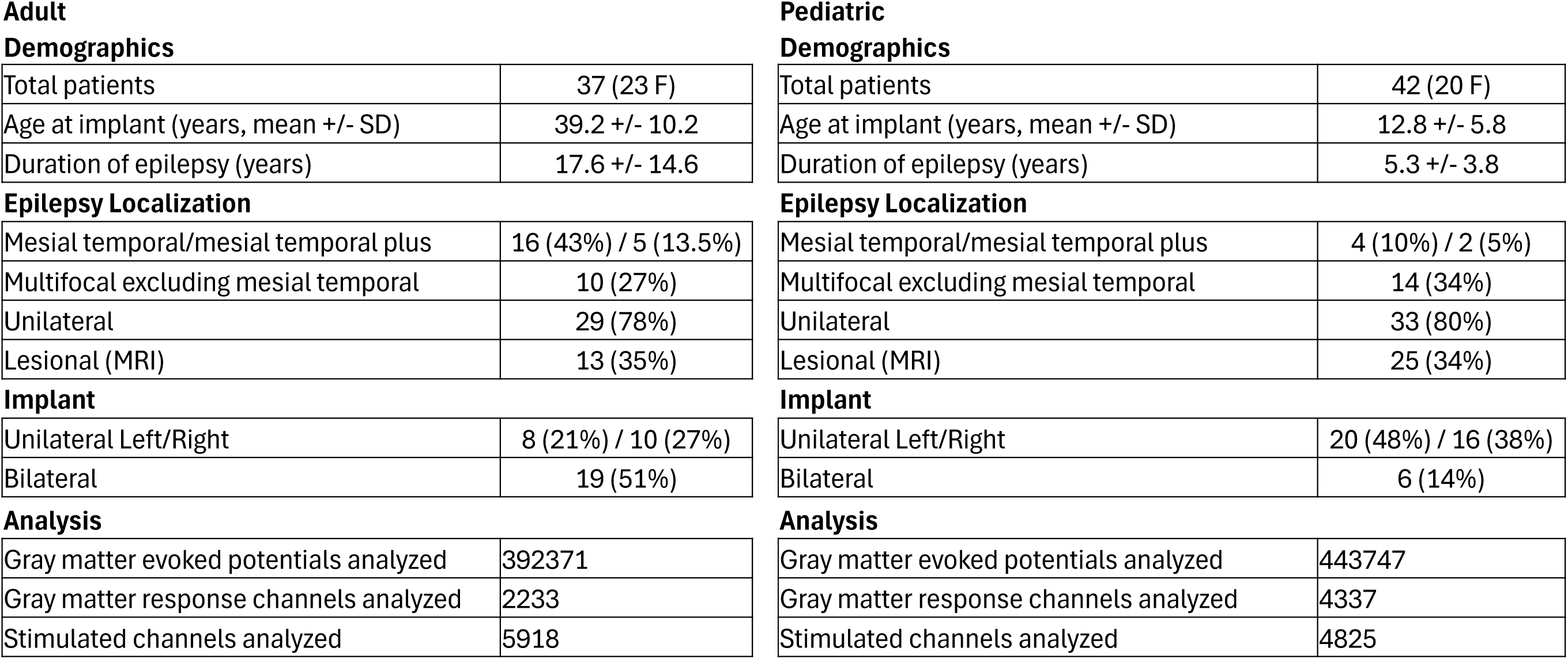
Study population.

Demographics
|  |  |
| --- | --- |
| Total patients | 37 (23 F) |
| Age at implant (years, mean +/- SD) | 39.2 +/- 10.2 |
| Duration of epilepsy (years) | 17.6 +/- 14.6 |
| Epilepsy Localization |  |
| Mesial temporal/mesial temporal plus | 16 (43%) / 5 (13.5%) |
| Multifocal excluding mesial temporal | 10 (27%) |
| Unilateral | 29 (78%) |
| Lesional (MRI) | 13 (35%) |
| Implant |  |
| Unilateral Left/Right | 8 (21%) / 10 (27%) |
| Bilateral | 19 (51%) |
| Analysis |  |
| Gray matter evoked potentials analyzed | 392371 |
| Gray matter response channels analyzed | 2233 |
| Stimulated channels analyzed | 5918 |
| Demographics |  |
| Total patients | 42 (20 F) |
| Age at implant (years, mean +/- SD) | 12.8 +/- 5.8 |
| Duration of epilepsy (years) | 5.3 +/- 3.8 |
| Epilepsy Localization |  |
| Mesial temporal/mesial temporal plus | 4 (10%) / 2 (5%) |
| Multifocal excluding mesial temporal | 14 (34%) |
| Unilateral | 33 (80%) |
| Lesional (MRI) | 25 (34%) |
| Implant |  |
| Unilateral Left/Right | 20 (48%) / 16 (38%) |
| Bilateral | 6 (14%) |
| Analysis |  |
| Gray matter evoked potentials analyzed | 443747 |
| Gray matter response channels analyzed | 4337 |
| Stimulated channels analyzed | 4825 |

### Analysis: Directed connectivity from evoked responses

Evoked responses produced by stimulation are inherently directional, since they are caused by perturbation at the site of stimulation. To analyze the stimulation-evoked response data, we followed our previous methods.^15^ The machine annotations generated in the EEG by the Natus stimulator device were used to identify time periods of stimulation and the identity of stimulated channels. Within these periods, an algorithm^15^ identified the large periodic electrical artifacts caused by stimulation. These were used to time-lock the signals recorded following each stimulation, generating an (average) evoked potential. The signals were averaged across individual stimulations following rejection of any epochs with signal greater than 1000 μV suggesting artifact; signals which are not phase locked to stimulation are thus minimized. These averaged waveforms were used for further analysis. As our primary goal was to broadly capture the magnitude of the evoked response from stimulation, and we preferred to avoid assumptions regarding any particular wave shape (see reference ^20^ for a discussion of this reasoning), we opted to calculate the root mean squared (RMS) deviation of the averaged waveform from the time window starting at 15 ms post-stimulation (to avoid effects of the stimulation artifact) extending to 250 ms post-stimulation. To control for any baseline differences between the signals recorded at different bipolar channels, we then scaled this quantity for each bipolar derivation, dividing the post-stimulation RMS by the RMS of the average signal from 100 ms to 15 ms immediately prior to stimulation. Reasoning that any evoked response should exhibit a higher magnitude signal compared to the pre-stimulation baseline, we restricted our analyses to signals with a scaled RMS greater than 1. Unless otherwise specified, subsequent evoked response analyses utilize only signals recorded from GM.

### Analysis: Designation of SOZ at stimulation and response sites

We used the clinical team’s notes to label each bipolar channel according to whether or not it was within the SOZ. If either contact within a bipolar pair was in the SOZ, the bipolar channel was considered to be within the SOZ. The evoked potential data is intrinsically two dimensional (see Fig 1), with one dimension representing stimulation site and the other representing response site. Each evoked response can be labeled as SOZ or not according to where it was recorded (response sites – either resp-SOZ or resp-nonSOZ) or according to where the provoking stimulation occurred (stim-SOZ or stim-nonSOZ). Such labeling will be employed throughout this manuscript, and is visualized in Figure 1.

**Figure 1:**
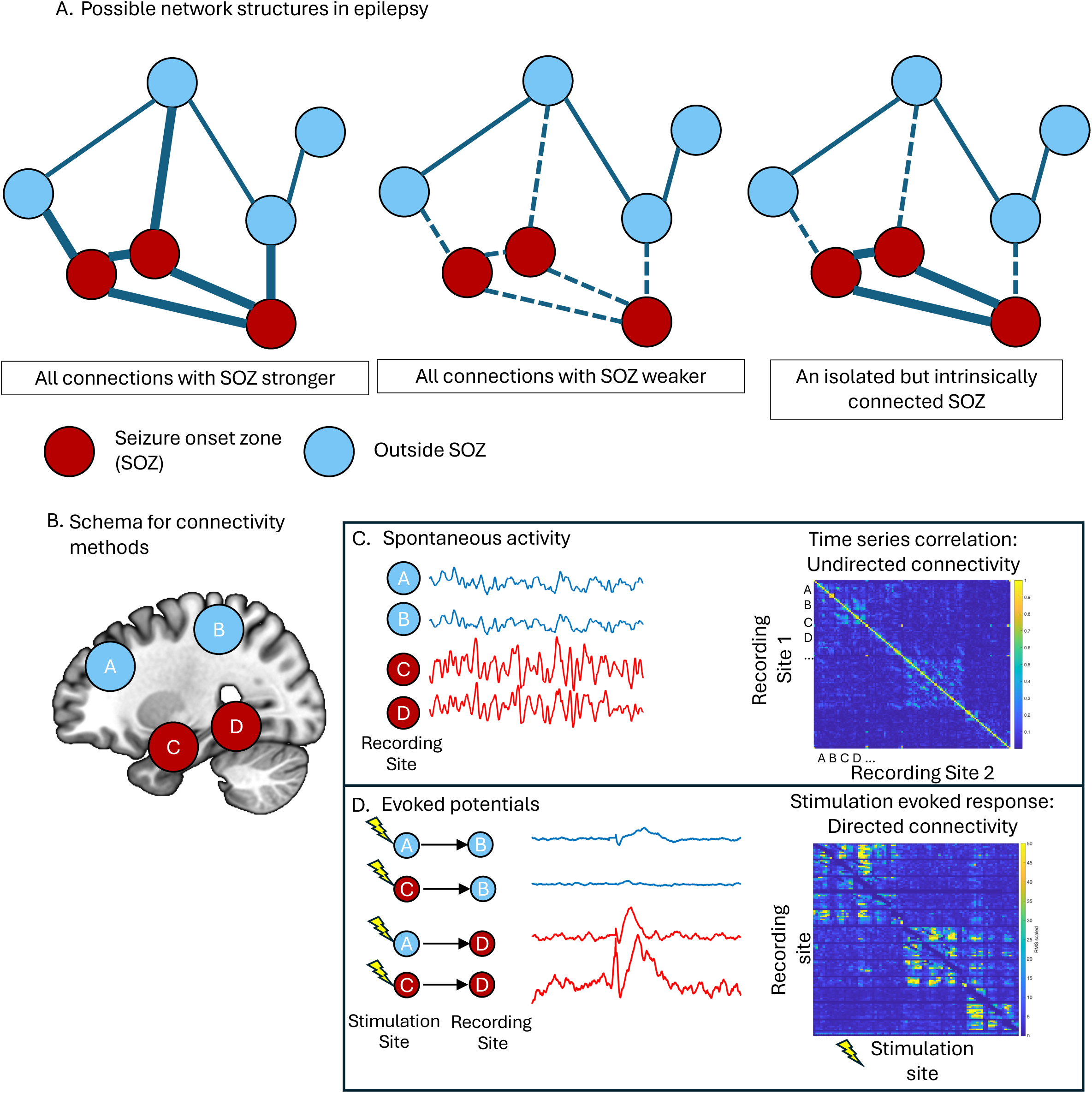
Visual hypotheses and methods. A. Simplified possible brain networks in epilepsy are shown, with thicker lines indicating stronger connections and dashed lines indicating weaker ones. B. On the left is a template with regions color-coded by whether they are in the seizure onset zone (SOZ - red) or not (blue), as used in this figure and throughout the other figures. C. Correlations between spontaneous activity recorded at different sites can be computed, resulting in a symmetric matrix of correlation values that represents undirected connectivity. D. In contrast, directed connectivity can be estimated by stimulating one site and recording the response at a second site. The evoked waveforms represent connectivity from the stimulation site to the recording site, and can be measured by calculating the root mean square of the evoked response wave form. Performing this calculation at each recording site, then doing the same for stimulation at multiple stimulation sites, results in a non-symmetric, directed connectivity matrix as shown on the bottom right.

### Analysis: Control for confounding effect of distance

Recording closer to a stimulation site typically yields larger evoked responses.^21,22^ Electrode contacts also tend to be spaced closer in the SOZ due to the clinical reasoning dictating their placement, thus raising the possibility of spuriously large signals simply due to the tighter spacing in the SOZ (see supplementary figure 1A for an illustration of this finding in our data set).^8^ Therefore, we first built a regression model using the distance between stimulation and response sites to predict the RMS values calculated from all non-SOZ recording sites located in GM. Similar to prior work,^16,23^ we utilized a rational polynomial of the form:

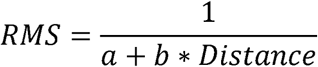

Where *RMS* is the measure of the evoked response magnitude and *Distance* is the pairwise distance between the stimulation and recording sites for that evoked response. This regression explained a large portion of the variance (96% in adult, 94% in pediatric data) in the non-SOZ RMS values, motivating the need to control for inter-contact distance. By evaluating the right-hand side of the equation for each evoked potential, an estimate of the distance-related portion of RMS can be obtained – subtracting this from the recorded RMS values yields the residual error term (RMS residuals). While the parameters used for this regression were estimated from non-SOZ tissue only, the resultant equation was applied to all channels. An identical procedure was performed for baseline connectivity values to remove/minimize the confounding effect of distance (Supplementary Figure 1).

### Analysis: Regressions

Linear mixed-effects regression models were utilized to analyze the relationship between physiologic signals and predictors of interest.

For analysis of baseline (undirected) connectivity, we labeled each correlation value according to where the pair of bipolar derivations was located with respect to the SOZ – either both outside the SOZ, both inside the SOZ, or one within and one outside the SOZ. The mean values for these categories are plotted with between-subjects standard error bars for visualization (Figure 2a), but regression analyses were utilized for statistical testing. Regression analyses permit modeling not only the fixed effects of interest (e.g., SOZ), but also potentially confounding covariates (interchannel distance); additionally, random effects were used to account for the measurements’ variability within subjects and within stimulation/recording channels. The resultant regression equation took the form:

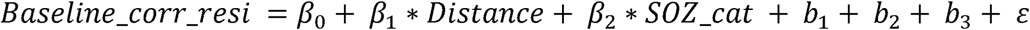

**Figure 2:**
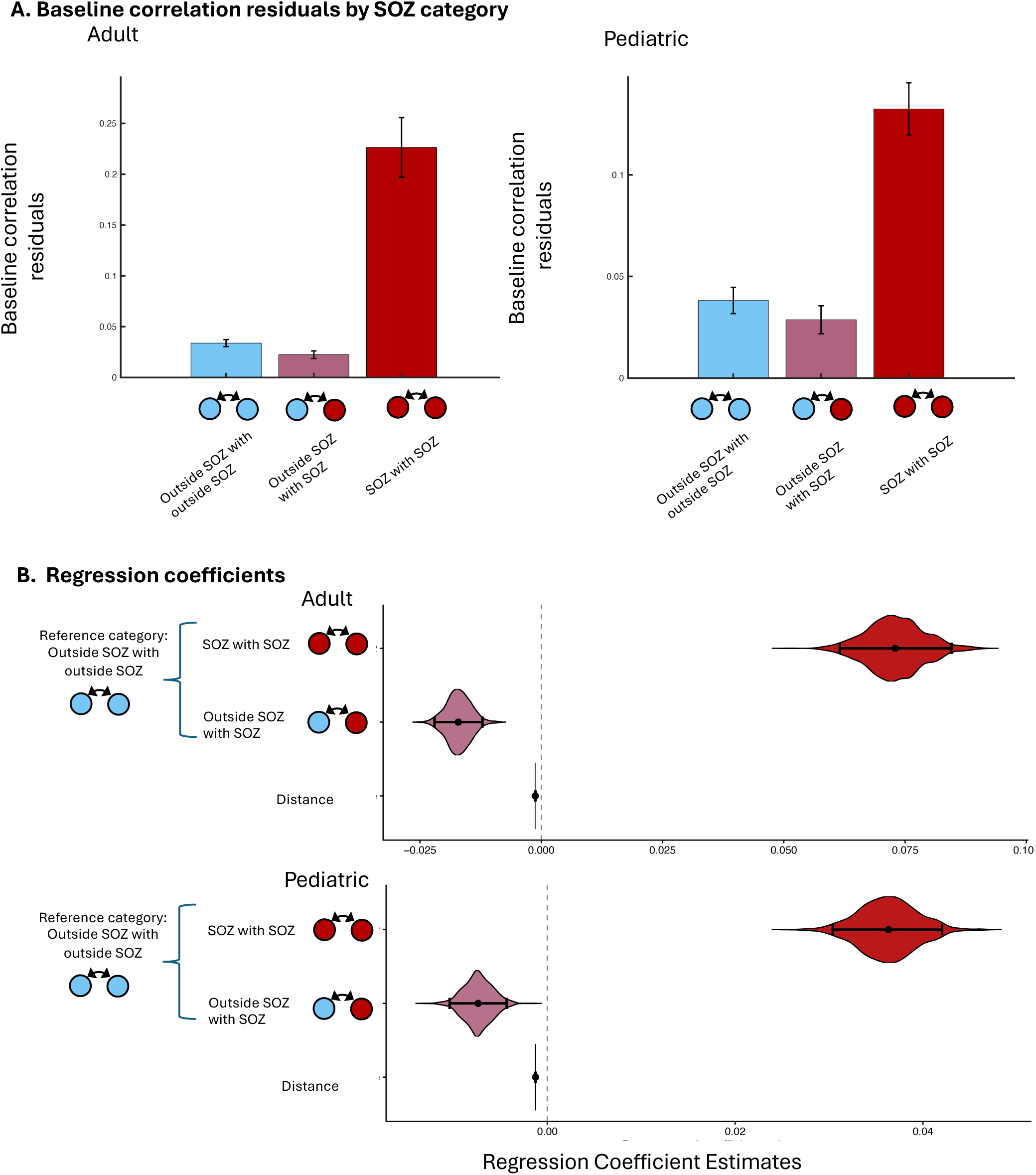
Undirected connectivity from correlations of baseline spontaneous activity. A. Mean values of adult (left) and pediatric (right) baseline correlations plotted according to whether each bipolar derivation is located within the SOZ or outside the SOZ. Error bars denote between-subject standard errors and are shown for illustrative purposes only; significance testing is performed with regression analyses, depicted in B, which account for between and within subjects errors (see Methods). B. Violin plots of regression coefficient estimates for SOZ and distance predicting baseline correlation residuals for adult (top) and pediatric (bottom) samples. Shown are the fixed effects coefficients for pairwise distance, and a categorical variable for SOZ of the channel pairs (the reference condition is outside SOZ with outside SOZ). Estimated regression coefficients are indicated by the black dots, with the violin shapes indicating the parametric bootstrapping distribution and the braces denoting 95% confidence intervals.

These variables are defined as follows: *Baseline_corr_resi* is the baseline correlation value between each pair of channels after the removal of distance as described above; *Distance* is the pairwise distance between bipolar channels; SOZ_cat is a categorical variable denoting whether neither, one, or both of the channels is in SOZ; the *β_n_* are the model intercept and regression coefficients associated with the fixed effects; b_1_ is the random intercept of channel 1 in the bipolar pair, b_2_ is the random intercept of channel 2, and b_3_ is the random intercept of subjects; and ε is the error term. To account for the possibility that a residual linear distance effect remained, we included pairwise *Distance* as a separate covariate.

For evoked responses, we anticipated that there might be differential effects of recording and stimulating within the SOZ, so we modeled these categorical predictors separately. The resultant regression equation took the form:

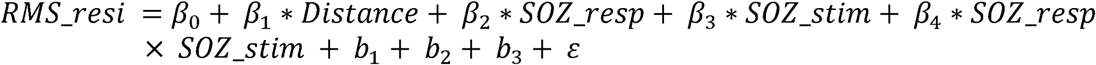

Here, *RMS_resi* represents the RMS of the evoked response after the modeled effect of distance was removed. *SOZ_resp* and *SOZ_stim* are binary variables denoting whether the response and stimulation site, respectively, are in the SOZ, modeled as fixed effects with an interaction term, with corresponding fixed effects coefficients *β_n_*. We included individual stimulation sites (corresponding to each column illustrated in Figure 1D) and individual response sites (corresponding to each row illustrated in Figure 1D) as random effects b_1_ and b_2_, respectively. Individual subjects were also modeled with a random effect term b_3_, and again *Distance* was included to account for any remaining variance related to pairwise stim/resp channel distance. Again, *ε* is the error term.

Similar regression analyses were performed for tissue type, labeling each data point according to whether the channel pair (for baseline correlation analyses) or stimulation/response channels were located within gray matter (GM) or white matter (WM).

Finally, in order to better assess statistical significance in the setting of many (>100,000) observations, which has the potential to create artificially low p-values and overly tight confidence intervals, we estimated confidence intervals by performing a parametric bootstrapping procedure with 1000 simulated datasets using the R function *bootmer*.^24^ The number of iterations was chosen based on pilot analyses, which showed stability in the estimates across multiple bootstrapping runs. The 95% confidence intervals were obtained by identifying the values at the 2.5^th^ and 97.5^th^ percentiles. Two-tailed p-values were estimated by calculating the proportion of the distribution greater/less than zero for positive/negative effects, respectively, then doubling it; in the event that a distribution’s range did not include zero, a p-value was assigned using the number of iterations (two-tailed p<0.002 for 1000 iterations).^25(pp181–182)^

## Results

For adult patients, the age (mean +/- standard deviation) was 39.2 +/- 10.2, and for the pediatric patients, the mean age was 12.8 +/- 5.8. The groups had differing distributions of epilepsy subtypes and duration of epilepsy (**Table 1**). A total of 392,371 evoked potential averaged waveforms were analyzed for the adult data set and an additional 443,747 in the pediatric data sets.

### Baseline connectivity

First, we analyzed connectivity by computing pairwise correlations for spontaneous neural activity recorded prior to stimulation. After removing the confounding effect of distance, we performed regression analyses on the residual correlation values, using the SOZ label of the bipolar channels as a categorical predictor for the pairwise correlation values. For both adults and pediatric patients, the intrinsic connectivity within SOZ was higher than between regions outside SOZ (Fig 2; regression coefficients in supplemental Table 1: adult β=0.073, 95% CI [0.062, 0.085], pediatric β=0.036 [0.030, 0.042], both p < 0.002). The connectivity between SOZ and non-SOZ, in contrast, was reduced for both data sets (adult β=-0.0170 [-0.022 to -0.012], pediatric β=-0.007 [-0.010, -0.004], both p < 0.002) compared to channel pairs located entirely outside SOZ. These results indicate that baseline connectivity is elevated between pairs of channels when both are located within the SOZ, but reduced for channel pairs comprising a SOZ and a non-SOZ channel, all relative to the baseline condition of pairs of non-SOZ channels.

### Stimulation connectivity

Next, we examined the pattern of evoked potentials. After removing the modeled effect of distance as described above, we performed regression analyses using categorical labels for SOZ of the stimulation/recording channels to predict the evoked potential RMS residuals, including random effects for patient and each individual response/stimulation channel. In the adult group (Fig. 3), RMS residuals from stimulating outside the SOZ and recording within the SOZ (nonSOZ→SOZ: β=-0.858 [-1.435, -0.290], p<0.004) were smaller relative to the reference category of nonSOZ→nonSOZ. RMS residuals generated from stimulating within the SOZ while recording outside the SOZ (SOZ→nonSOZ) were also smaller (β=-1.299 [-1.939, - 0.616], p<0.002) relative to the same reference. However, there was an interaction such that both stimulating and recording within the SOZ yielded larger RMS residuals than the nonSOZ→nonSOZ reference (β=1.589 [0.837, 2.387], p<0.002).

**Figure 3:**
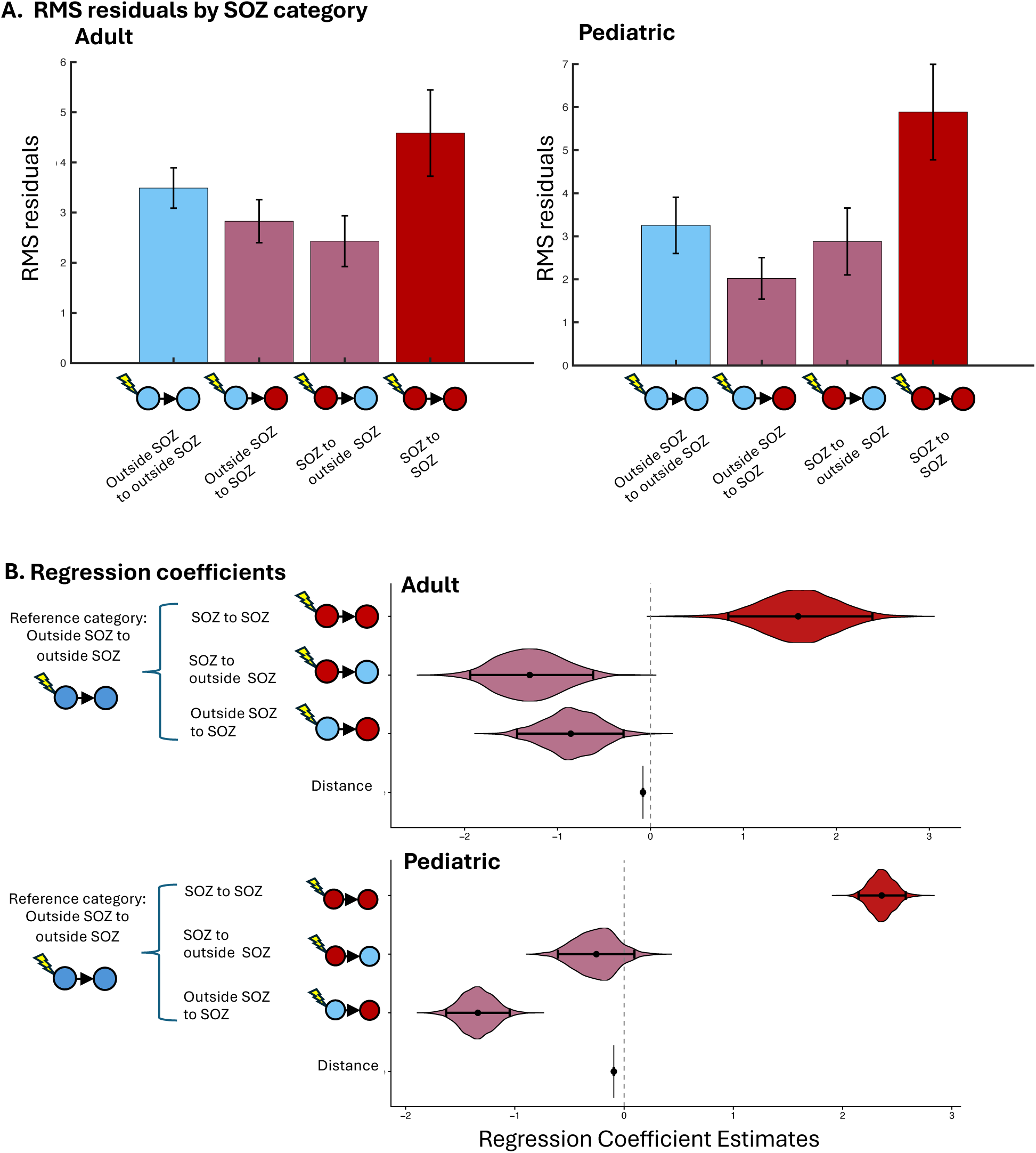
Directed connectivity from evoked responses. A. Mean values of adult (left) and pediatric (right) evoked potential RMS residuals, plotted according to whether each stimulation and response bipolar channel is located within the SOZ or outside the SOZ. Error bars denote between-subject standard errors and are shown for illustrative purposes only; significance testing is performed with regression analyses, depicted in B, which account for between and within subjects errors (see Methods). B. Coefficient estimates for SOZ and distance predicting evoked response RMS residuals for adult (top) and pediatric (bottom) samples; shown are fixed effects for pairwise distance between stimulating and response channels, categorical variables denoting whether the stimulating and response channels are located in the seizure onset zone, and an interaction term for the latter two variables (the reference condition is stimulating outside SOZ and recording outside SOZ). Estimated regression coefficients are indicated by the black dots, with the violin shapes indicating the parametric bootstrapping distribution and the braces denoting 95% confidence intervals.

For the pediatric data, the same connectivity pattern was seen for nonSOZ→SOZ (β=-1.338 [-1.628, -1.047], p<0.002) and for the interaction of SOZ→SOZ (β=2.36 [2.146, 2.577], p < 0.002), all relative to the same reference category of nonSOZ→nonSOZ. In the pediatric data set, however, the SOZ→nonSOZ effect remained numerically negative but was not significant (β=-0.254 [-0.606, -0.095], p=0.166 – see supplementary Table 2 for full regression results).

A supplemental analysis including baseline correlation residuals as a covariate revealed that, as expected, higher baseline correlations predicted higher evoked responses, and otherwise yielded similar results for the SOZ factors (Supplemental Fig 3), suggesting that the relationship between SOZ and evoked potentials remains when baseline connectivity is accounted for – although, as in the main analysis, non-significantly so in the pediatric data set for SOZ→nonSOZ connectivity. Next, as a control against the possibility that the initial calculation of residuals from the distance regression could have artificially reduced the signals in the SOZ, we repeated the regression models without the initial distance regression step, with similar results (see supplemental Figure 2 and supplemental Table 5).

These results overall agree with the baseline connectivity results, and suggest that stimulation connectivity is elevated between contacts within the SOZ, but reduced between SOZ and non-SOZ contacts.

### Effect of tissue type

Next, we aimed to understand how stimulation and recording in different tissue types might affect the baseline connectivity and evoked responses in our data. Here, again utilizing residual RMS values after removing the main effect of distance (as described in *Methods*), we computed a regression using only non-SOZ channels to understand the relationship between tissue type and baseline connectivity. These regression analyses revealed that for both adult and pediatric data sets, baseline correlation residuals differed amongst different tissue types, with pairs of GM channels showing reduced connectivity relative to WM channel pairs (adult β=-0.011 [-0.015, -0.006], p<0.002; pediatric β= -0.014 [-0.018, -0.010], p < 0.002). Pairs comprising a GM and a WM channel also showed reduced baseline connectivity compared to WM channel pairs (adult β=-0.012 [-0.014, -0.009], p<0.002; pediatric β=-0.014 [-0.018 to -0.010], p<0.002; see Figure 4a and 4b, Supplementary Table 3).

**Figure 4:**
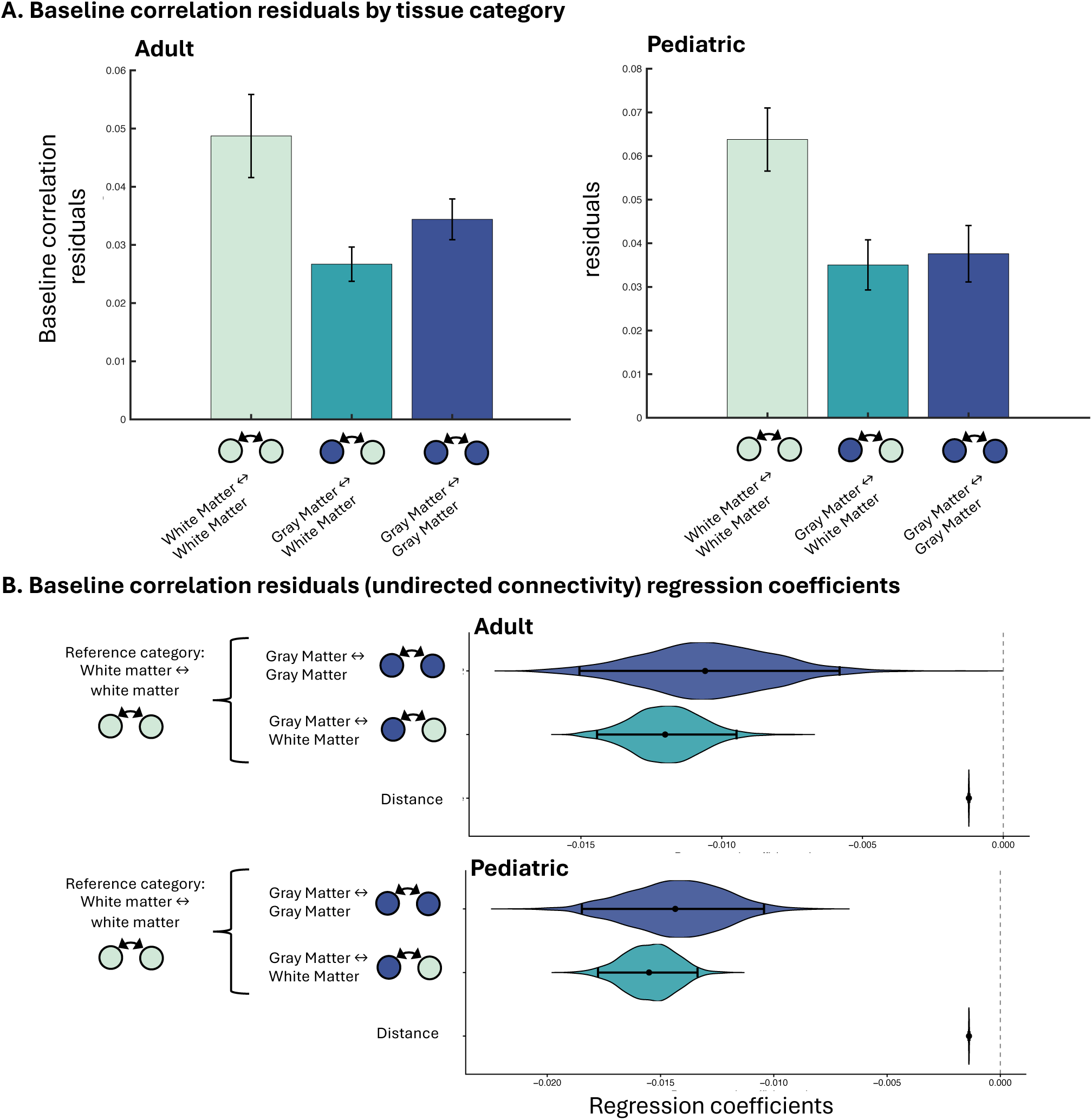
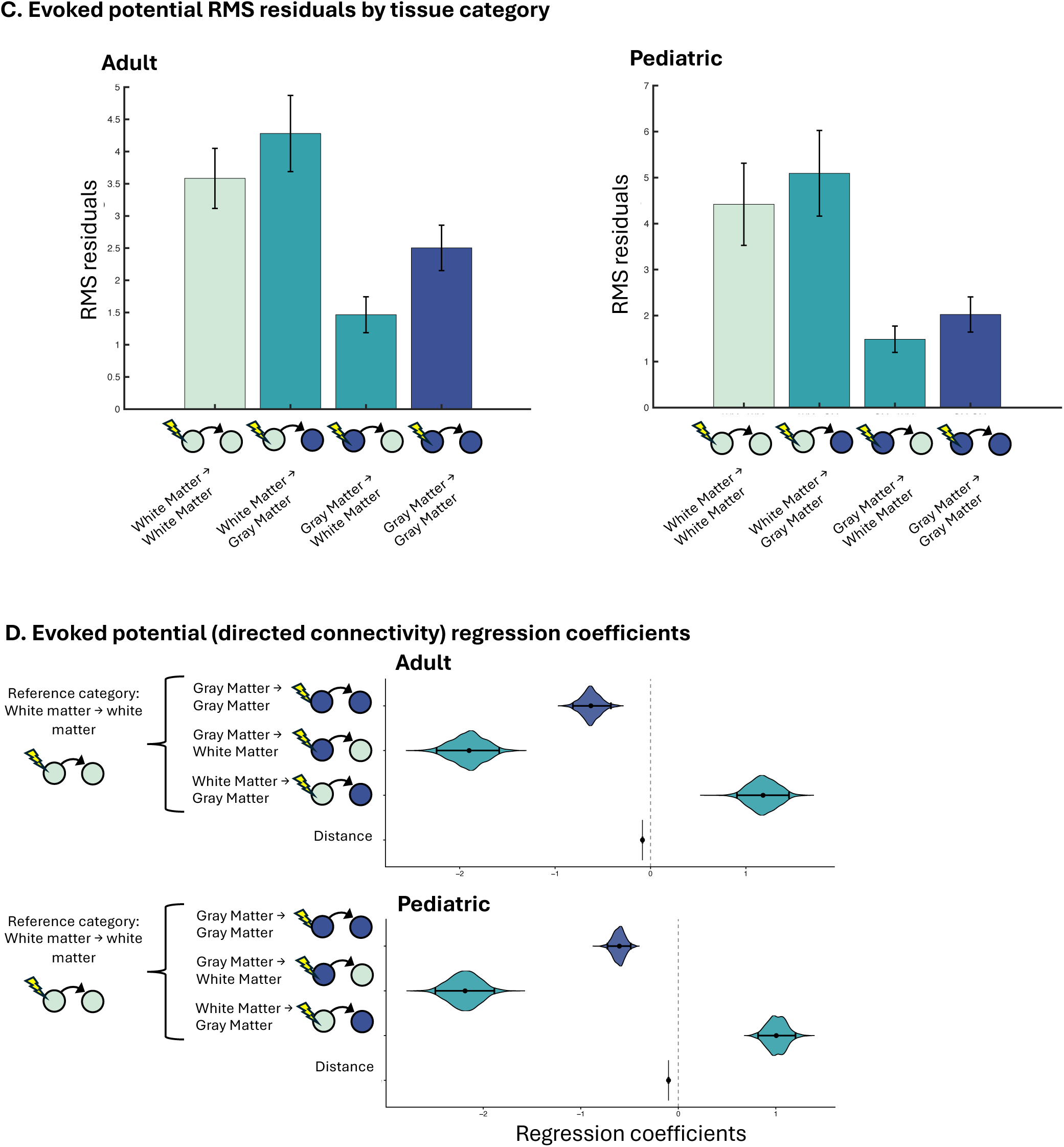
Effect of tissue type on connectivity measures. A. Average baseline between-channel correlations split according to type of tissue in which channels are located (open circles indicate white matter, and gray circles indicate gray matter), for only non-SOZ contacts. Error bars denote standard errors across subjects, and are illustrative only; significance testing is performed via regression analyses. B. Regression coefficients from regression predicting baseline connectivity from tissue type (gray or white matter) of each bipolar channel, with white matter to white matter connectivity as the reference condition. Estimated regression coefficients are indicated by the black dots, with the violin shapes indicating the parametric bootstrapping distribution and the braces denoting 95% confidence intervals. Only non-SOZ channels were used in this analysis. C. Evoked potential RMS plotted according to tissue type of stimulation and response site, for only non-SOZ contacts. Error bars denote between-subject standard error only for visualization; significance testing is performed with regression analyses.

We also compared evoked potential RMS residuals by tissue type. As illustrated in Figure 4c and 4d, compared with the baseline category of both stimulating and recording within WM, stimulating in WM while recording within GM yielded larger RMS residuals (adult β=1.18 [0.906, 1.451], p<0.002); however, stimulating GM, regardless of the recording site, yielded smaller RMS residuals (β=-1.901 [-2.243, -1.586], p<0.002 for GM→WM, and β=-0.625 [-0.814, -0.415], p<0.002 for GM→GM – see supplementary Table 4). For the pediatric data set, an identical pattern of results was obtained (β=-1.005 [0.818, 1.201], p<0.002 for WM→GM, β=-2.186 [-2.491, -1.887], p<0.002 for GM→WM, and β=-0.604 [-0.726, -0.487], p<0.002 for GM→GM). Thus, stimulating in the white matter tends to produce higher responses, but responses are higher when recorded in gray matter.

## Discussion

How connectivity is altered within the epileptogenic network remains debated, limiting the use of network measures as biomarkers to guide surgery. To address this, we quantified connectivity using both resting state intracranial EEG and stimulation-evoked responses. Across modalities and in cohorts from an adult and a pediatric center, we observed a consistent pattern: connectivity within the SOZ was increased, whereas connections from SOZ to non-SOZ regions were reduced (with one exception – see below). These findings support a model of an *internally hyperconnected but isolated EZ*—increased connectivity within the SOZ accompanied by disconnection from the broader network.

### Resting connectivity is increased within SOZ but reduced between SOZ and non-SOZ tissue

This result, consistent across the adult and pediatric data sets, aligns with prior work from intracranial EEG suggesting higher interictal connectivity within the SOZ in patients who had good outcomes following epilepsy surgery^9,26,27^. On the other hand, connectivity between SOZ electrodes and regions outside the SOZ was reduced, which echoes prior work which identified reduced large scale connectivity in focal epilepsy^6,8,14,27,28^. These resting state findings support the hypothesis of an isolated but internally hyperconnected EZ.

### Stimulation connectivity is also increased within the SOZ but reduced between SOZ and non-SOZ tissue

Mirroring our resting state findings, evoked potentials were largest when both the stimulating and recording electrodes were in the SOZ, but reduced when only one of the pair (stimulator or recorder) was in the SOZ. This SOZ↔SOZ elevation is consistent with prior stimulation studies.^11–14,29^ By contrast, cross-boundary pairs (SOZ↔non-SOZ) showed attenuated responses compared to non-SOZ↔non-SOZ (only trending for the pediatric SOZ→non-SOZ condition), supporting a functionally isolated EZ. These findings persisted when baseline connectivity was included in the regression models, suggesting that the relationship between SOZ and stimulation-derived connectivity is not fully accounted for by baseline connectivity. Notably, we did not observe an effect consistent with the recently described interictal suppression hypothesis—which predicts stronger directed influence from non-SOZ→SOZ and weaker SOZ→non-SOZ.^30^ We observed a trend in this direction in adult patients, but the opposite direction was seen in pediatric patients (Fig. 3B); indeed, the SOZ→non-SOZ evoked potentials were not significantly smaller than baseline in the pediatric group. However, Johnson et al. reported frequency-specific effects; our RMS metric may blur band-limited asymmetries, suggesting that band-resolved analyses could reveal directionality not captured here. It is also possible that different epileptic pathologies in children versus adults lead to interictal suppression by non-SOZ sites only in the latter.^31^

### Distance and tissue type affects connectivity and stimulation responses

We replicated prior observations that resting connectivity is higher between nearby contacts^32^ and stimulation responses are higher in electrode contacts located near the stimulation site.^21,33^ Because clinical electrode implantation is typically denser around the suspected EZ, this can bias connectivity estimates toward higher values within the SOZ.^8,16^ We observed consistent findings in our connectivity analyses both before and after adjusting for inter-electrode distance, but it is possible that a residual distance confound persists and partially explains the finding of higher connectivity within the SOZ. Future work comparing homologous left–right structures within a homogeneous syndrome (e.g., temporal lobe epilepsy) could further control for spatial sampling biases.

Stimulation responses were lower when *recording* in white matter, consistent with the understanding that cortical pyramidal neurons are the primary generators of local field potentials.^34^ In contrast, *stimulating* white matter produced larger CCEP responses. This has been reported by other groups, particularly with greater long-distance responses to white-matter stimulation.^35^ A likely mechanistic explanation is that white matter stimulation directly depolarizes long-range axons, facilitating distant signal propagation, whereas gray matter stimulation engages both excitatory pyramidal cells and inhibitory interneurons,^36^ potentially yielding a smaller net effect. Baseline connectivity values varied between gray matter and white matter, which further justifies the logic of restricting the SOZ analyses to gray matter only. Finally, stimulating gray matter produced *smaller* gray matter evoked potentials relative to stimulating in white matter; this finding argues against the possibility that the heightened evoked potentials seen in SOZ→SOZ pairs could be due to higher propensity for SOZ channels to be in gray matter.

*Potential mechanisms underlying an isolated but intrinsically hyperconnected EZ* Human microelectrode recordings reveal a sharp boundary between a hypersynchronous ictal core and a low-firing penumbra, consistent with inhibitory restraint that limits spread.^37,38^ Two-photon Ca²⁺ imaging in mice shows interneuron engagement around the focus, supporting a narrow inhibitory ring that can suppress propagation.^39,40^ This core–surround architecture naturally yields strong within-SOZ coupling but weak SOZ↔non-SOZ communication—a locally reinforced yet globally isolated EZ. Extending these findings across human epileptic pathologies with higher-density microelectrodes, and in animal models with cell-type–resolved Ca²⁺ imaging/perturbations, will test the scope and mechanisms of this inhibitory surround.

### Consistent patterns of EZ connectivity across ages

The differences in EEG resting and perturbed connectivity across the lifespan are poorly understood. A recent study of CCEPs across the lifespan revealed a gradual decrease in conduction delays with age until at least 30 years old, suggesting that the speed of brain communication increases throughout childhood and as a young adult.^41^ We found that the pattern of connectivity with respect to the SOZ remained consistent between our two data sets, despite differences in age and epilepsy syndromes. However, we restricted our analysis to the *amplitude*, rather than the *speed*, of evoked responses. Future analyses of latency could further probe the change in conduction across the lifespan.

### Implications for surgical epilepsy

An EZ that is locally hyperconnected yet globally isolated is not optimally captured by a single scalar per contact (e.g., row/column averages of an adjacency matrix). Such summaries mix strong within-SOZ edges with weak SOZ↔non-SOZ edges, diluting the signal. Translational work should instead target pattern-level contrasts, such as within-vs between-region connectivity, variance/heterogeneity across a contact’s edges, or module/atlas-based metrics that quantify boundary strength. Optimizing and validating these pattern-sensitive measures could yield a practical EZ biomarker measurable at rest, which could be used to dramatically shorten iEEG evaluations.

### Limitations

The strategy of utilizing 1 Hz stimulation without jittered timing raises the possibility that entrainment of neural signals may have affected our results.^42,43^ Also, while the collection of all data in a single session should reduce within-subject variability, this also prevents us from testing or controlling for the impact of cognitive state or medications.^44–47^ Our limited sample size with incomplete spatial sampling also precludes us from comparing responses according to anatomical location of stimulation/response site or according to epilepsy localization. Finally, the use of all subjects regardless of outcome is a significant caveat, as we cannot confirm that the SOZ reflects the EZ.

## Conclusions

Our convergent interictal correlational and stimulation-based findings seen in both a pediatric and adult epilepsy cohort point to an EZ that is locally hyperconnected yet globally isolated. This organization helps reconcile prior cross-modal inconsistencies and aligns with basic science demonstrating a hypersynchronous ictal core surrounded by inhibitory restraint. For translational impact, network-based biomarkers should prioritize pattern-sensitive metrics that emphasize within-SOZ vs. SOZ↔non-SOZ contrasts rather than simple per-contact averages. If successful, these approaches could deliver a resting-state biomarker that shortens invasive monitoring and improves surgical decision-making.

## Data Availability

All neurophysiologic data is available at ieeg.org. All analytical codes are located at https://github.com/erinconrad/CCEPS.

https://ieeg.org

https://github.com/erinconrad/CCEPS

## Supplemental Figure Captions

Supplemental Table 1: Regression results for the pairwise (by bipolar derivation) baseline correlations (BL_mean_corr), with predictor variables of pairwise distance and category of channel pair (SOZ-Out represents bipolar derivation pairs with one bipolar derivation in the SOZ and one outside the SOZ; SOZ-SOZ represents a correlation between two bipolar derivations both within the SOZ). The beta coefficient from the model fit to the actual data is shown for each fixed effect, along with 95% confidence intervals and distribution of estimates from the parametric bootstrapping analysis. For the purposes of this regression, correlations between two bipolar derivations both outside the SOZ serve as the baseline category against which the other two are compared.

Supplemental Table 2: Regression results for the residuals of RMS of the stimulation evoked responses, with predictor variables of pairwise distance and category of channel pair. Estimate refers to the regression beta coefficient, with corresponding confidence intervals and p-values obtained from parametric bootstrapping. In this regression, stimulating nonSOZ and recording in nonSOZ serves as the baseline category against which the others are compared. Below are plotted coefficient estimates from similar regressions including baseline connectivity as a co-predictor of RMS.

Supplemental Table 3: Regression results for the pairwise (by channel pair) baseline correlations, with predictor variables of pairwise distance and category of channel pair (WM=white matter, GM=gray matter). WM-GM represents channel pairs with channel in GM and one in WM; GM-GM represents a correlation between two channels both within GM. The beta coefficient from the model fit to the actual data is shown for each fixed effect, along with 95% confidence intervals and distribution of estimates from the parametric bootstrapping analysis. For the purposes of this regression, correlations between two channels both in WM serve as the baseline category against which the other two are compared.

Supplemental Table 4: Shown are the regression coefficient estimates for the analysis of RMS residuals of the stimulation evoked responses, after the initial step of regressing out the effect of distance. The predictor variables of model intercept, pairwise distance, and category of channel pair with respect to tissue type (WM=white matter, GM=grey matter) are shown; these analyses additionally included random effects for individual subject, stimulation channel, and response channel. The beta coefficient from the model fit to the actual data is shown for each fixed effect, along with 95% confidence intervals and distribution of estimates from the parametric bootstrapping analysis. In this regression, stimulating WM and recording in WM serves as the baseline category against which the others are compared.

Supplemental Table 5: Shown are the regression coefficient estimates for the analysis of RMS of the stimulation evoked responses, omitting the initial step of regressing out the effect of distance. The predictor variables of model intercept, pairwise distance, category of channel pair are shown; these analyses additionally included random effects for individual subject, stimulation channel, and response channel. The beta coefficient from the model fit to the actual data is shown for each fixed effect, along with 95% confidence intervals from the parametric bootstrapping analysis. In this regression, both stimulating and recording in nonSOZ serves as the baseline category against which the others are compared.

Supplemental Figure 1: Evoked potential RMS values from adult data (left) (and pediatric data (right) plotted against pairwise distance between stimulation and response channels. Histograms adjacent to the y- and x-axis show the distributions of RMS and pairwise distances, respectively. Superimposed in green is the line of best fit as described in *Methods*, with the equation y = p1/(x+q1), where y represents the RMS values, x represents the pairwise distances, and p1/q1 are constants. For adults, these values are p1=566.46 and q1=106.43. On the right is the equivalent plot for the pediatric data, with constants p1=404.95 and q1=59.6

Supplemental Figure 2: Regression results for the residuals of RMS of the stimulation evoked responses, with predictor variables of pairwise distance, category of channel pair, and baseline correlation from each channel pair. Estimate refers to the median regression beta coefficient from bootstrapping over the residuals for 1000 iterations, with corresponding confidence intervals and p-values for these coefficient estimates. These analyses additionally included random effects for individual subject, stimulation channel, and response channel. Estimated regression coefficients are indicated by the black dots, with the violin shapes indicating the parametric bootstrapping distribution and the braces denoting 95% confidence intervals. In this regression, stimulating nonSOZ and recording in nonSOZ serves as the baseline category against which the others are compared.

Supplemental Figure 3: Shown are the regression coefficient estimates for the analysis of RMS of the stimulation evoked responses, omitting the initial step of regressing out the effect of distance. The predictor variables of pairwise distance, category of channel pair, and baseline correlation from each channel pair are shown; these analyses additionally included random effects for individual subject, stimulation channel, and response channel. Estimated regression coefficients are indicated by the black dots, with the violin shapes indicating the parametric bootstrapping distribution and the braces denoting 95% confidence intervals. In this regression, stimulating nonSOZ and recording in nonSOZ serves as the baseline category against which the others are compared.

