## Supplementary figures and images for "Intrinsic and extrinsic connectivity of the seizure onset zone at rest and during stimulation"

### Supplemental Figure 1

#### Supplement Figure 1: Distance regressions

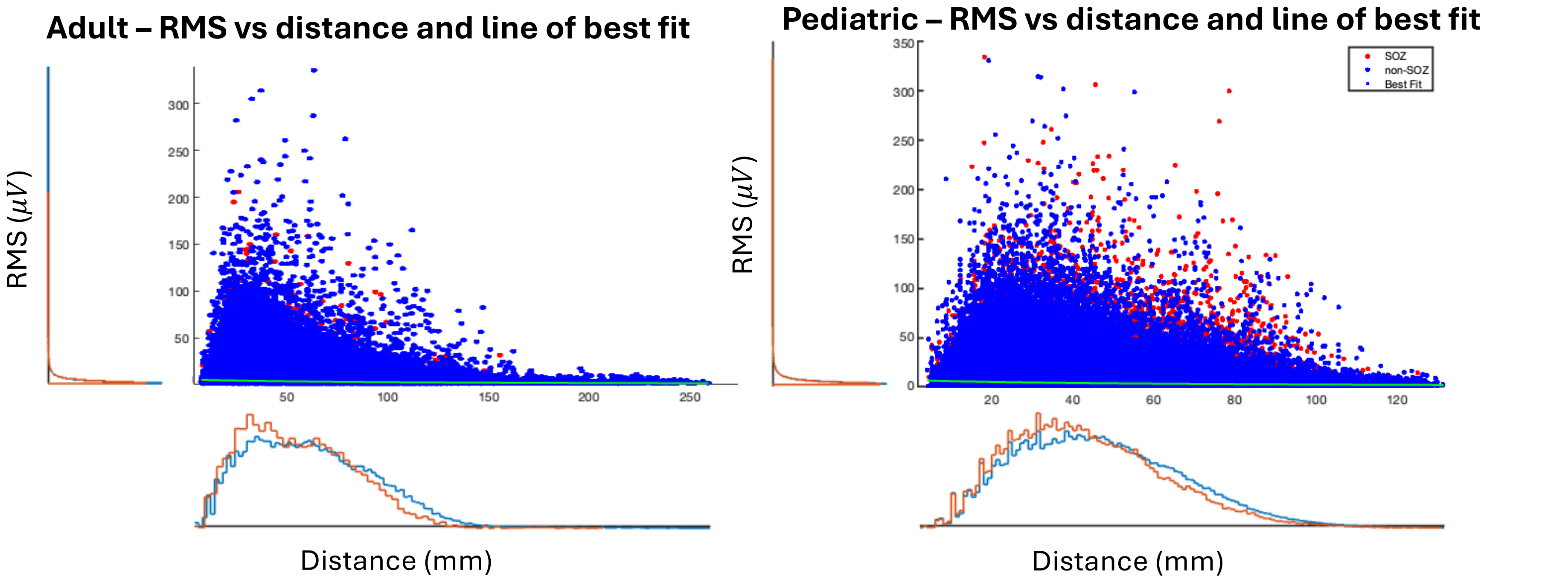

### Supplemental Figure 2

#### Supplement Figure 2: Directed connectivity regression including baseline connectivity

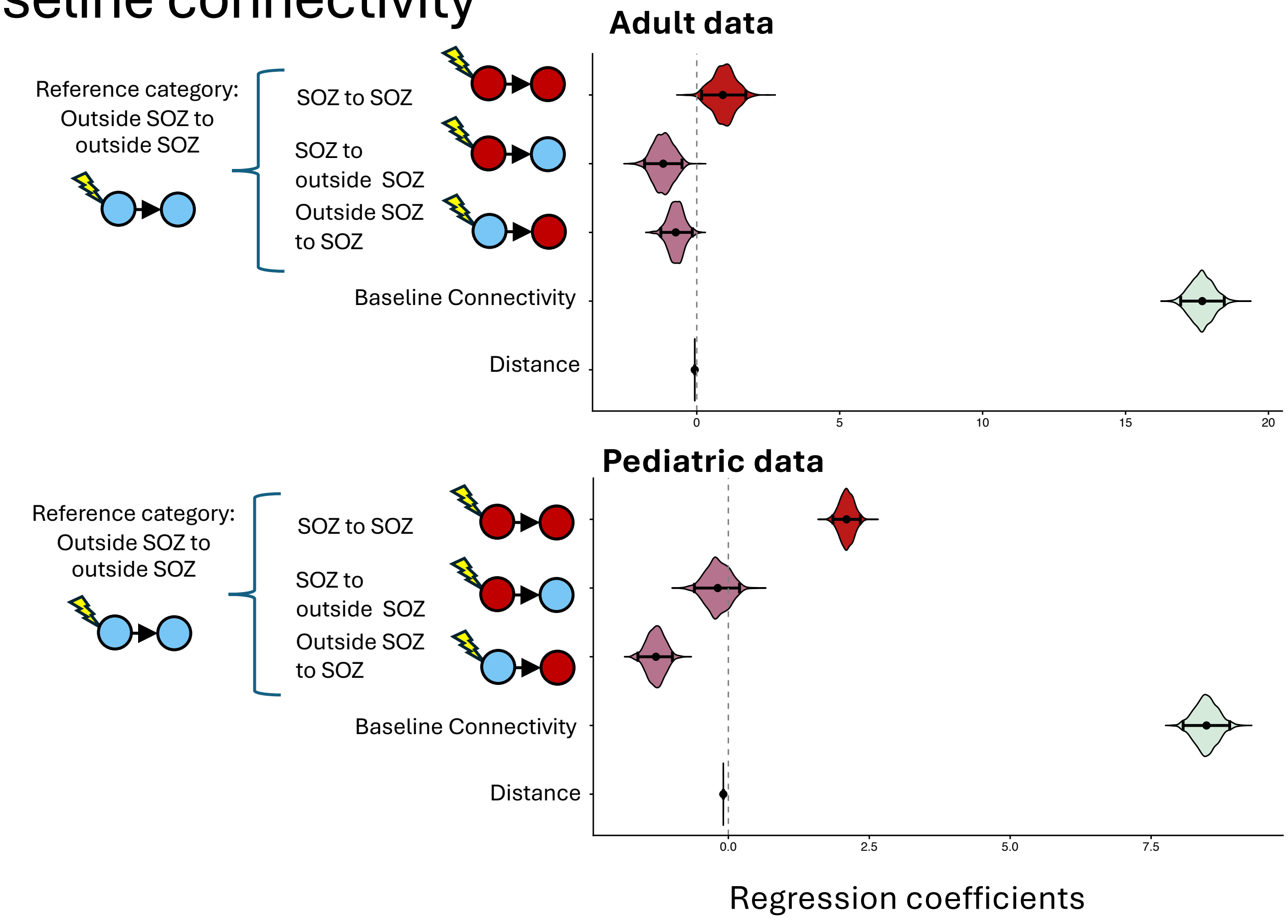
