## Supplemental Figure 3 for "Intrinsic and extrinsic connectivity of the seizure onset zone at rest and during stimulation"

### Supplement Figure 3: Regressions for directed connectivity from stimulation-evoked potentials, omitting initial distance correction step

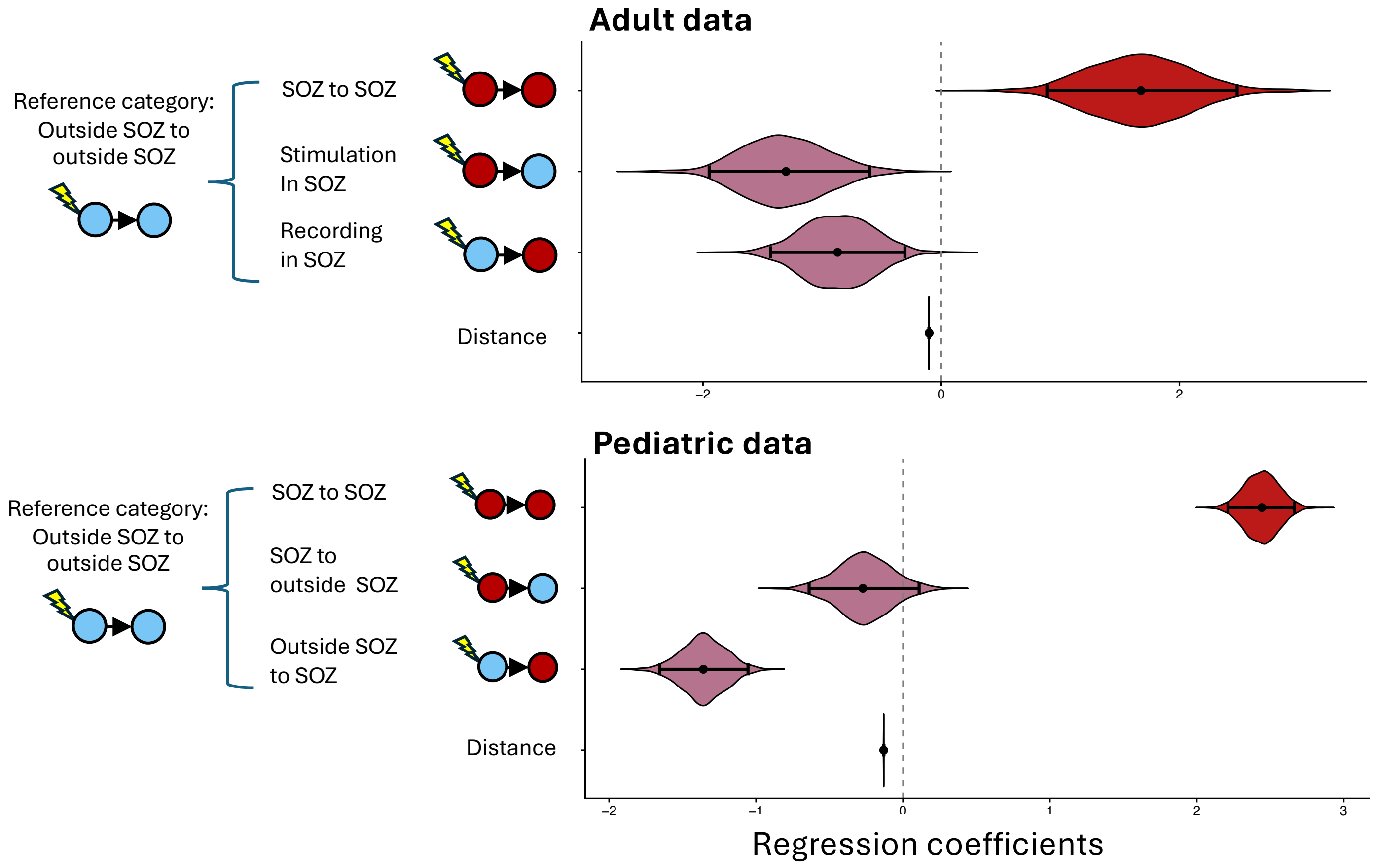
