## Supplemental Table 1 for "Intrinsic and extrinsic connectivity of the seizure onset zone at rest and during stimulation"

### Supplement Table 1: Regression tables for undirected baseline connectivity for SOZ

#### Adults

Baseline_correlation_residuals ~ 1 + Distance + SOZ_category+ (1 | bipolar_chan1) + (1 | bipolar_chan2) + (1 | subject)

| Name | Estimate | Confidence Interval | pValue |
|---|---|---|---|
| (Intercept) | 0.108 | [0.101, 0.115] | <0.002 |
| Distance | -0.001 | [-0.001,-0.001] | <0.002 |
| SOZ-Out | -0.017 | [-0.022, -0.012] | <0.002 |
| SOZ - SOZ | 0.073 | [0.062, 0.085] | <0.002 |

#### Peds

Baseline_correlation_residuals ~ 1 + Distance + SOZ_category+ (1 | bipolar_chan1) + (1 | bipolar_chan2) + (1 | subject)

| Name | Estimate | Confidence Interval | pValue |
|---|---|---|---|
| (Intercept) | 0.100 | [0.087, 0.113] | <0.002 |
| Distance | -0.001 | [-0.001,-0.001] | <0.002 |
| SOZ-Out | -0.007 | [-0.010, -0.004] | <0.002 |
| SOZ - SOZ | 0.036 | [0.030, 0.042] | <0.002 |
