## Supplemental Table 2 for "Intrinsic and extrinsic connectivity of the seizure onset zone at rest and during stimulation"

### Supplement Table 2: Regressions for directed connectivity from stimulation-evoked potentials for SOZ

#### Adult

RMS_residuals ~ 1 + Distance + + SOZ_resp + SOZ_stim + SOZ_resp*SOZ_stim + (1 | resp_chan) + (1 | stim_chan) + (1 | subject)

| Name | Estimate | Confidence Interval | pValue |
|---|---|---|---|
| (Intercept) | 8.100 | [7.284, 8.833] | <0.002 |
| Distance | -0.081 | [-0.083,-0.079] | <0.002 |
| nonSOZ -> SOZ | -0.858 | [-1.435, -0.290] | 0.004 |
| SOZ -> nonSOZ | -1.299 | [-1.939, -0.616] | <0.002 |
| SOZ -> SOZ | 1.589 | [0.837, 2.387] | <0.002 |

#### Pediatric

RMS_residuals ~ 1 + Distance + + SOZ_resp + SOZ_stim + SOZ_resp*SOZ_stim + (1 | resp_chan) + (1 | stim_chan) + (1 | subject)

| Name | Estimate | Confidence Interval | pValue |
|---|---|---|---|
| (Intercept) | 7.132 | [6.17, 8.063] | <0.002 |
| Distance | -0.094 | [-0.096,-0.092] | <0.002 |
| nonSOZ -> SOZ | -1.338 | [-1.628, -1.047] | <0.002 |
| SOZ -> nonSOZ | -0.254 | [-0.606, 0.095] | 0.166 |
| SOZ -> SOZ | 2.358 | [2.146, 2.577] | <0.002 |

#### Adult – including baseline

RMS_residuals ~ 1 + Distance + SOZ_resp*SOZ_stim + Baseline_Correlation_Residuals + (1 | resp_chan) + (1 | stim_chan) + (1 | subject)

| Name | Estimate | Confidence Interval | pValue |
|---|---|---|---|
| (Intercept) | 7.113 | [6.368, 7.883] | <0.002 |
| Distance | -0.071 | [-0.072, -0.069] | <0.002 |
| Baseline_Correlation_Residuals | 17.683 | [16.928, 18.453] | <0.002 |
| nonSOZ -> SOZ | -0.735 | [-1.257, -0.155] | <0.01 |
| SOZ -> nonSOZ | -1.173 | [-1.825, -0.516] | 0.004 |
| SOZ -> SOZ | 0.913 | [0.170, 1.718] | 0.02 |

#### Pediatric – including baseline

RMS_residuals ~ 1 + Distance + SOZ_resp*SOZ_stim + Baseline_Correlation_Residuals + (1 | resp_chan) + (1 | stim_chan) + (1 | subject)

| Name | Estimate | Confidence Interval | pValue |
|---|---|---|---|
| (Intercept) | 6.832 | [5.759, 7.953] | <0.002 |
| Distance | -0.089 | [-0.091,-0.087] | <0.002 |
| Baseline_Correlation_Residuals | 8.48 | [8.068, 8.894] | <0.002 |
| nonSOZ -> SOZ | -1.285 | [-1.604, -0.993] | <0.002 |
| SOZ -> nonSOZ | -0.189 | [-0.602, 0.198] | 0.394 |
| SOZ -> SOZ | 2.097 | [1.856, 2.341] | <0.002 |
