## Supplemental Table 3 for "Intrinsic and extrinsic connectivity of the seizure onset zone at rest and during stimulation"

### Supplement Table 3: Regressions for undirected baseline connectivity for tissue type

#### Adult

Baseline_correlation_residuals ~ 1 + Distance + GM_category + (1 | bipolar_chan1) + (1 | bipolar_chan2) + (1 | subject)

| Name | Estimate | Confidence Interval | pValue |
|---|---|---|---|
| (Intercept) | 0.121 | [0.114, 0.128] | <0.002 |
| Distance | -0.001 | [-0.001,-0.0001] | <0.002 |
| WM - GM | -0.012 | [-0.014, -0.009] | <0.002 |
| GM - GM | -0.011 | [-0.015, -0.006] | <0.002 |

#### Pediatric

Baseline_correlation_residuals ~ 1 + Distance + GM_category + (1 | bipolar_chan1) + (1 | bipolar_chan2) + (1 | subject)

| Name | Estimate | Confidence Interval | pValue |
|---|---|---|---|
| (Intercept) | 0.122 | [0.109, 0.135] | <0.002 |
| Distance | -0.001 | [-0.001,-0.001] | <0.002 |
| WM - GM | -0.016 | [-0.018, -0.013] | <0.002 |
| GM - GM | -0.014 | [-0.018, -0.010] | <0.002 |
