## Supplemental Table 4 for "Intrinsic and extrinsic connectivity of the seizure onset zone at rest and during stimulation"

### Supplement Table 4: Regressions for directed connectivity from stimulation-evoked potentials for tissue type

#### Adult

RMS_residuals ~ 1 + Distance + GM_resp + GM_stim + GM_resp*GM_stim + (1 | resp_chan) + (1 | stim_chan) + (1 | subject)

| Name | Estimate | Confidence Interval | pValue |
|---|---|---|---|
| (Intercept) | 8.476 | [7.638, 9.223] | <0.002 |
| Distance | -0.084 | [-0.086,-0.082] | <0.002 |
| WM -> GM | 1.180 | [0.906, 1.451] | <0.002 |
| GM -> WM | -1.901 | [-2.243, -1.586] | <0.002 |
| GM -> GM | -0.625 | [-0.814, -0.415] | <0.002 |

#### Pediatric

RMS_residuals ~ 1 + Distance + GM_resp + GM_stim + GM_resp*GM_stim + (1 | resp_chan) + (1 | stim_chan) + (1 | subject)

| Name | Estimate | Confidence Interval | pValue |
|---|---|---|---|
| (Intercept) | 8.447 | [7.477, 9.494] | <0.002 |
| Distance | -0.100 | [-0.102,-0.098] | <0.002 |
| WM -> GM | 1.005 | [0.818, 1.201] | <0.002 |
| GM -> WM | -2.186 | [-2.491, -1.887] | <0.002 |
| GM -> GM | -0.604 | [-0.726, -0.487] | <0.002 |
