## Supplemental Table 5 for "Intrinsic and extrinsic connectivity of the seizure onset zone at rest and during stimulation"

### Supplement Table 5: Regressions for directed connectivity from stimulation-evoked potentials, omitting initial distance correction step

#### Adult

RMS ~ 1 + Distance + + SOZ_resp + SOZ_stim + SOZ_resp*SOZ_stim + (1 | resp_chan) + (1 | stim_chan) + (1 | subject)

| Name | Estimate | Confidence Interval | pValue |
|---|---|---|---|
| (Intercept) | 12.818 | [12.009, 13.649] | <0.002 |
| Distance | -0.101 | [-0.103,-0.099] | <0.002 |
| nonSOZ -> SOZ | -0.870 | [-1.433, -0.304] | <0.004 |
| SOZ -> nonSOZ | -1.303 | [-1.948, -0.599] | <0.002 |
| SOZ -> SOZ | 1.677 | [0.887, 2.484] | <0.002 |

#### Pediatric

RMS ~ 1 + Distance + + SOZ_resp + SOZ_stim + SOZ_resp*SOZ_stim + (1 | resp_chan) + (1 | stim_chan) + (1 | subject)

| Name | Estimate | Confidence Interval | pValue |
|---|---|---|---|
| (Intercept) | 12.824 | [11.871, 13.782] | <0.002 |
| Distance | -0.131 | [-0.133,-0.130] | <0.002 |
| nonSOZ -> SOZ | -1.358 | [-1.657, -1.055] | <0.002 |
| SOZ -> nonSOZ | -0.273 | [-0.639, 0.110] | 0.172 |
| SOZ -> SOZ | 2.442 | [2.212, 2.665] | <0.002 |
